# Brain Network Connectivity in Adolescents At Risk for Alcohol Use Problems

**DOI:** 10.1101/2020.10.22.20217976

**Authors:** Simon T. E. Baker, Murat Yücel, Alex Fornito, Andrew Zalesky, Sarah Whittle, Nicholas B. Allen, Dan I. Lubman

**Affiliations:** Turning Point, Eastern Health and Eastern Health Clinical School, Monash University, Victoria, Australia; Brain and Mental Health Laboratory, Monash Institute of Cognitive and Clinical Neurosciences, Monash University, Victoria, Australia; Melbourne Neuropsychiatry Centre, Department of Psychiatry, University of Melbourne and Melbourne Health, Victoria, Australia; Melbourne School of Engineering, University of Melbourne, Victoria, Australia; Melbourne School of Psychological Sciences, University of Melbourne, Victoria, Australia; Orygen, The National Centre of Excellence in Youth Mental Health, Victoria, Australia

**Keywords:** adolescence, alcohol, graph theory, risk, structural connectivity, white matter

## Abstract

Imaging studies of young people with a family history of alcohol use disorder (AUD) have found structural and/or functional differences within and between anatomically distributed and functionally specialised systems throughout the brain. Differences in brain connectivity among adolescents with a family history of AUD may account for the increased risk of later alcohol use problems; however, to date, no prospective studies have directly examined this hypothesis across the entire connectome in a regionally unbiased way. Our analysis included 52 adolescents (*M*_age_ = 16.5 years ± 0.6 *SD*) assessed with diffusion-weighted magnetic resonance imaging, of whom 20 had a family history of AUD and 32 did not. All participants were followed-up 2.3 years later and completed a questionnaire measuring past year alcohol use and alcohol-related harms. Subject-specific connectomic maps of structural connectivity were constructed using two parcellation schemes (82-node anatomical and 530-node random) and five measures of connectivity weight (streamline count, fractional anisotropy, mean diffusivity, axial diffusivity, and radial diffusivity), and a connectome-wide network-based statistic analysis was used to compare group differences at each and every connection between adolescents with and without a family history of AUD. Baseline connectivity measures did not differentiate these groups, and we did not find an association between baseline connectivity measures and alcohol outcomes at follow-up. These findings suggest that atypical inter-regional structural connectivity may not contribute to the risk of developing alcohol use problems in this particular age group, or during this particular period of development.

---

A family history of alcohol use disorder (AUD) is associated with an earlier age of beginning to drink alcohol and, in subsequent years, a higher prevalence of alcohol use problems, including AUD (Chassin, Pitts, DeLucia, & Todd, 1999; Hill, Shen, Lowers, & Locke, 2000; Sher, Walitzer, Wood, & Brent, 1991). The underlying causes of this increase in risk remain unclear, but studies with a primary focus on elucidating its neural basis have provided specific clues. In brain imaging studies, young people with a family history of AUD show structural and/or functional differences within and between anatomically distributed and functionally specialised systems (e.g., frontal, parietal, temporal, and subcortical brain regions) compared to those with no such family history (see the following reviews: Cservenka, 2016; Cservenka, Alarcon, Jones, & Nagel, 2015). Such findings suggest that dysconnectivity between brain regions may be a neural correlate of increased risk for the later development of alcohol use problems (Cservenka et al., 2015).

Magnetic resonance imaging (MRI) studies of differences in brain connectivity between young people with and without a family history of AUD have identified aberrant functional connectivity between the striatum and frontal and parietal cortices (Cservenka, Casimo, Fair, & Nagel, 2014; Spadoni, Simmons, Yang, & Tapert, 2013; Weiland et al., 2013; Wetherill et al., 2012), the amygdala and frontal cortex (Cservenka, Fair, & Nagel, 2014), and the cerebellum and frontal cortex (Herting, Fair, & Nagel, 2011), as well as microstructural white matter abnormalities along frontostriatal (Squeglia et al., 2015) and several other white matter tracts (Acheson et al., 2014; Herting, Schwartz, Mitchell, & Nagel, 2010; Squeglia, Jacobus, Brumback, Meloy, & Tapert, 2014) in those with a family history of AUD. These findings are consistent with the hypothesis that pre-existing, heritable patterns of brain connectivity may be an important determinant of one’s ability to regulate behaviours such as alcohol consumption (Clark, Thatcher, & Tapert, 2008). However, a prospective analysis of the relationship between brain connectivity and subsequent alcohol use in young people with a family history of AUD is lacking.

The aim of this study was to examine the possibility that atypical structural connectivity increases the risk of developing alcohol use problems, especially in those with a family history of such problems. To address this aim in a comprehensive and regionally unbiased way, diffusion-weighted MRI was performed in 52 adolescents (20 with a family history of AUD and 32 with no such family history) at the age of 16.5 years. Structural connectivity was modelled as a graph of nodes (brain regions) connected by edges (white matter tracts reconstructed by tractography). A connectome-wide analysis (Fornito, Zalesky, & Breakspear, 2015; Zalesky, Fornito, & Bullmore, 2010) was then performed to test for group differences at each and every pairwise connection between adolescents with and without a family history of AUD. An additional analysis was conducted to examine the association between patterns of structural connectivity and alcohol consumption, as well as alcohol-related harms, 2.3 years after the MRI scans were acquired. Based on the previous literature, we hypothesised that adolescents with a family history of AUD would show a distributed pattern of reduced structural connectivity compared to adolescents with no such family history. We also expected that this reduction of structural connectivity would correlate with subsequent levels of alcohol consumption and alcohol-related harms at follow-up.

## Method

### Participants

Participants were recruited as part of a longitudinal research project investigating biological, psychological, and social risk factors for psychopathology (see Whittle et al., 2014; Whittle et al., 2008). From among this community sample, we selected participants who had completed diffusion- and T1-weighted MRI sequences during the mid-adolescent wave of data collection (*M*_age_ = 16.5 years ± 0.5 *SD*). Participants were excluded if they had a major medical or neurological condition, contraindication related to medication, neurological or incidental radiological abnormality detected as part of routine MRI screening by a hospital radiologist, or history of head injury resulting in loss of consciousness. Participants were also excluded if they or a biological parent had a lifetime history of psychotic disorder (schizophrenia or bipolar I), or if their family history of psychiatric disorder was not known. Based on these criteria, and after quality control of MRI data, a final sample of 52 adolescents was selected (*M*_age_ = 16.5 years ± 0.6 *SD*). These adolescents were assigned to one of two groups on the basis of whether or not they had a biological parent retrospectively diagnosed with AUD, leaving 20 (38.5%) adolescents with a family history of AUD and 32 (61.5%) adolescents with no such family history. No adolescent had a family history of any substance use disorder (SUD) other than AUD.

Written informed consent was obtained from all adolescents and their parent or guardian in accordance with the guidelines from local research and ethics committees.

### Family History

Biological parents were assessed for lifetime history of AUD using the Structured Clinical Interview for DSM Disorders (First, Spitzer, Gibbon, & Williams, 1994). If one parent was unavailable, the available parent was invited to provide information on the unavailable parent in accordance with the Family History Research Diagnostic Criteria (Andreasen, Endicott, Spitzer, & Winokur, 1977).

### Alcohol Use and its Associated Harms

At baseline and at 2-year follow-up, the Youth Risk Behavior Surveillance System survey (Centers for Disease Control and Prevention, 2004) was used to assess patterns of alcohol and other drug use. Alcohol consumption was assessed with the question “During [your life / the past 12 months], on how many days have you had at least one drink of alcohol?” Use of other drugs (e.g., cannabis/marijuana, ecstasy/MDMA, methamphetamines, and heroin) was assessed with a series of questions asking “During [your life / the past 30 days], how many times have you used [the drug]?” Alcohol-related harms were assessed with the question “In the past 12 months, did your use of alcohol ever cause you to (a) get so drunk you were sick or passed out; (b) have trouble at home, work, or school; (c) get injured or have an accident; (d) become violent and get into a fight; (e) have sex with someone which you later regretted; (f) get in trouble with the police; (g) be unable to remember what happened the night before; (h) be asked to leave a party, pub, or club because you were drunk; (i) feel you were not able to stop drinking once you started; (j) feel irritable or depressed when it wasn’t available” (Little et al., 2013). Participants were instructed to circle yes or no for each of the 10 problems listed. Affirmative responses were summed to create a continuous measure of the number of alcohol-related harms experienced.

### MRI Acquisition

MRI was performed on a 3 Tesla system (MAGNETOM Trio, A Tim System, Siemens Medical Solutions) at the Royal Children’s Hospital, Melbourne, and included the following acquisitions: (a) high angular resolution diffusion imaging (slice thickness = 2.3 mm; repetition time = 7,300 msec; echo time = 104 msec; field of view = 240 mm^2^; image matrix = 104 × 104; voxel size = 2.3 mm^3^), comprised of 60 gradient-weighted (b-value = 3,000 s/mm^2^) volumes interspersed with seven T2-weighted (i.e., b-value = 0) volumes; and (b) high-resolution T1-weighted structural imaging (repetition time = 1,900 msec; echo time = 2.24 msec; flip angle = 9°; field of view = 230 mm^2^; image matrix = 256 × 256; voxel size = 0.9 mm^3^), with the volume comprising 176 contiguous 0.9 mm slices.

### MRI Processing and Connectivity Mapping

MRI data processing and structural connectivity mapping were conducted using well-validated analysis tools, including FreeSurfer (Fischl, 2012), FMRIB Software Library (FSL; Jenkinson, Beckmann, Behrens, Woolrich, & Smith, 2012), and MRtrix (Tournier, Calamante, & Connelly, 2012), as well as code developed in our laboratory.

Our pipeline included three major steps (see Baker et al., 2015, for further details). In the first step, using automated cortical reconstruction and subcortical segmentation tools available in FreeSurfer (Fischl, 2012), the T1-weighted volume was parcellated into 34 cortical (Desikan et al., 2006) and seven subcortical (Fischl et al., 2002) homologous regions in each hemisphere, totalling 82 distinct brain regions. Since parcellation can be somewhat arbitrary and can influence results (Fornito, Zalesky, & Bullmore, 2010; Zalesky et al., 2010b), results were verified using a random parcellation comprising 530 nodes (265 per hemisphere) of approximately equal volume, generated according to the methods described by Zalesky et al. (2010b) and Fornito et al. (2010; see also Fornito et al., 2011). The inverse transformation matrix from the linear alignment of an average of the seven T2-weighted (b-value = 0) volumes to the T1-weighted volume in Talairach space was applied to both parcellation schemes to align the parcellated regions to native diffusion space (Jenkinson, Bannister, Brady, & Smith, 2002).

In the second step, we performed constrained spherical deconvolution based probabilistic tractography to construct a “tractogram” comprising one million streamlines that collectively estimate the trajectories of white matter tracts (Tournier et al., 2012). This approach reconstructs tracts that correspond well with known white matter anatomy despite the challenges to tractography posed by crossing fibres (Jeurissen, Leemans, Jones, Tournier, & Sijbers, 2011; Tournier et al., 2012).

In the third and final step, each parcellation scheme was combined with the tractogram to produce a set of 10 connectivity matrices for each participant. We constructed an 82-node and a 530-node streamline count-weighted connectivity matrix by populating each [*i, j*] element of an *N* × *N* adjacency matrix, where *N* is equal to the number of nodes, with the number of streamlines intersecting brain region *i* and brain region *j*. A streamline intersecting more than two regions was assumed to connect the pair of regions that were maximally separated according to the Euclidean distance calculated using the centre of gravity of each region. To avoid the inclusion of spurious streamlines crossing the medial longitudinal fissure, interhemispheric streamlines were restricted to those with at least one point of propagation located in a corpus callosum mask. In addition to the streamline count, inter-regional connection weights were also quantified using four different scalar measures derived from the eigenvalues of the diffusion tensor that have been proposed to characterise the microstructural properties of white matter, including fractional anisotropy (FA), mean diffusivity (MD), axial diffusivity (AD), and radial diffusivity (RD; see Beaulieu, 2009). We constructed an 82-node and a 530-node FA-weighted connectivity matrix by populating each [*i, j*] element of an *N* × *N* adjacency matrix with a mean FA value, calculated as FA averaged over all voxels intersected by at least one of the set of streamlines connecting brain regions *i* and *j*. Thus, the edge weights of the FA-weighted connectivity matrices represented the mean FA of connections between brain regions. This procedure was repeated for MD, AD, and RD. Our pipeline yielded 10 different network representations of brain connectivity: (a) 82-node streamline count-weighted, (b) 82-node FA-weighted, (c) 82-node MD-weighted, (d) 82-node AD-weighted, (e) 82-node RD-weighted, (f) 530-node streamline count-weighted, (g) 530-node FA-weighted, (h) 530-node MD-weighted, (i) 530-node AD-weighted, and (j) 530-node RD-weighted.

### Statistical Analysis of Connectivity

To characterise group differences in baseline structural connectivity measures between adolescents with and without a family history of AUD, we computed a one-tailed independent-samples *t*-test at each edge of the connectivity matrix. For each of 3,321 (82-node) and 140,185 (530-node) unique connection estimates, the null hypothesis tested was equality in the average connectivity weight between both groups. Connections with a *t* statistic exceeding a primary threshold of *p* = .05, two-tailed, uncorrected, were admitted to a binarised matrix of suprathreshold connections for subsequent analysis with the Network Based Statistic (NBS; Zalesky et al., 2010a). The NBS is a validated nonparametric statistical procedure that, in the context of connectome-wide (brain network) analyses, offers greater statistical power than other methods commonly used to correct for multiple comparisons (e.g., Bonferroni and False Discovery Rate). This is because statistical inference with the NBS is performed at the level of interconnected subsets of edges, called connected components, rather than at each edge independently. Connected components were assigned a FWE corrected *p*-value using permutation testing (i.e., 10,000 random permutations of the original data). This *p*-value was used to determine whether the size of any observed components significantly exceeded levels expected by chance (FWE corrected, *p* < .05).

The NBS was also used to examine the prospective association between baseline structural connectivity measures and alcohol consumption as well as alcohol-related harms 2.3 years later. Within each individual connectivity matrix, the weight of each edge was corrected (demeaned) by subtracting from it the average weight of that edge across all subjects. The resulting connectivity matrices were regressed against demeaned alcohol data to test for an association between (a) baseline connectivity measures and the level of alcohol consumption in the 12 months prior to follow-up, and (b) baseline connectivity measures and the number of alcohol-related harms experienced in the 12 months prior to follow-up.

## Results

### Demographic and Behavioural Results

Participant characteristics and patterns of alcohol and other drug use are presented in Table 1. At the baseline assessment, which took place when the sample (*N* = 52) had a mean age of 16.5 years (± 0.6 *SD*, age range: 14.95–18.05 years), no group differences in age, sex, whole brain volume, Full Scale Intelligence Quotient (Wechsler, 2003), or handedness were observed between adolescents with (*n* = 20) and without (*n* = 32) a family history of AUD. Even so, we controlled for age in our NBS analyses of connectivity to account for any age-related differences. When assessed at baseline, 19 of 20 (95%) adolescents with a family history of AUD had consumed alcohol at least once in their lifetime, whereas 19 of 32 (59%) adolescents without a family history of AUD had consumed alcohol at least once in their lifetime. Accordingly, the group of adolescents with a family history of AUD had significantly more lifetime drinking episodes at baseline (range: 0–100 days) compared to the group of adolescents with no such family history (range: 0–30 days). Thus, we performed a supplementary series of NBS analyses to test for group differences in baseline structural connectivity measures between drinkers (*n* = 38) and non-drinkers (*n* = 14). Although this comparison indicated that there was no difference, we controlled for lifetime drinking history in our NBS analyses of connectivity at baseline, given the well-established effects of alcohol on brain structure and function. Adolescents with a family history of AUD also experienced more alcohol-related harms in the 12 months prior to baseline (14 [70%] reported no harm; 5 [25%] reported one harm; 1 [5%] reported two harms) compared to adolescents with no such family history (30 [94%] reported no harm; 2 [6%] reported one harm).

**Table 1.**
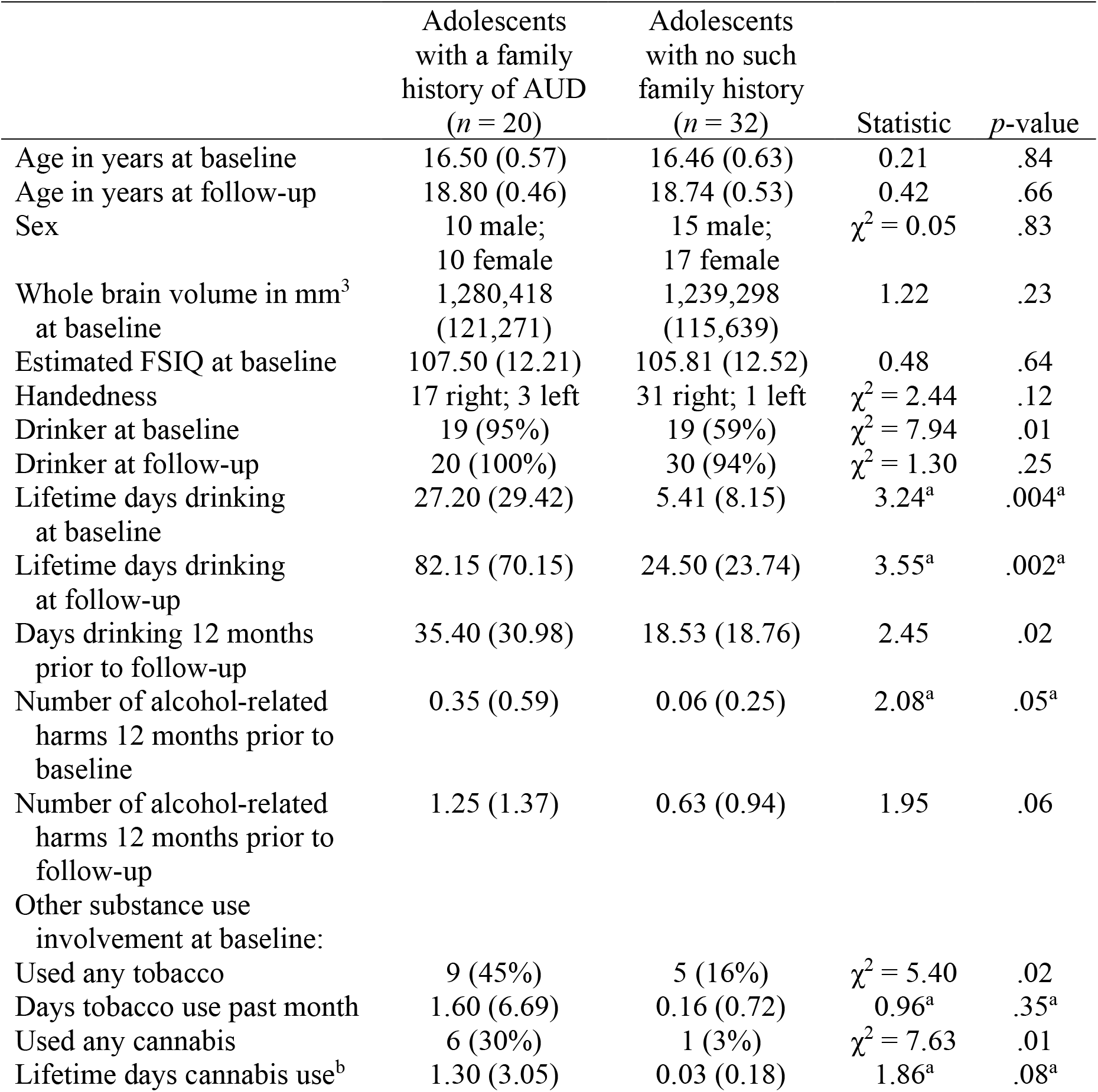
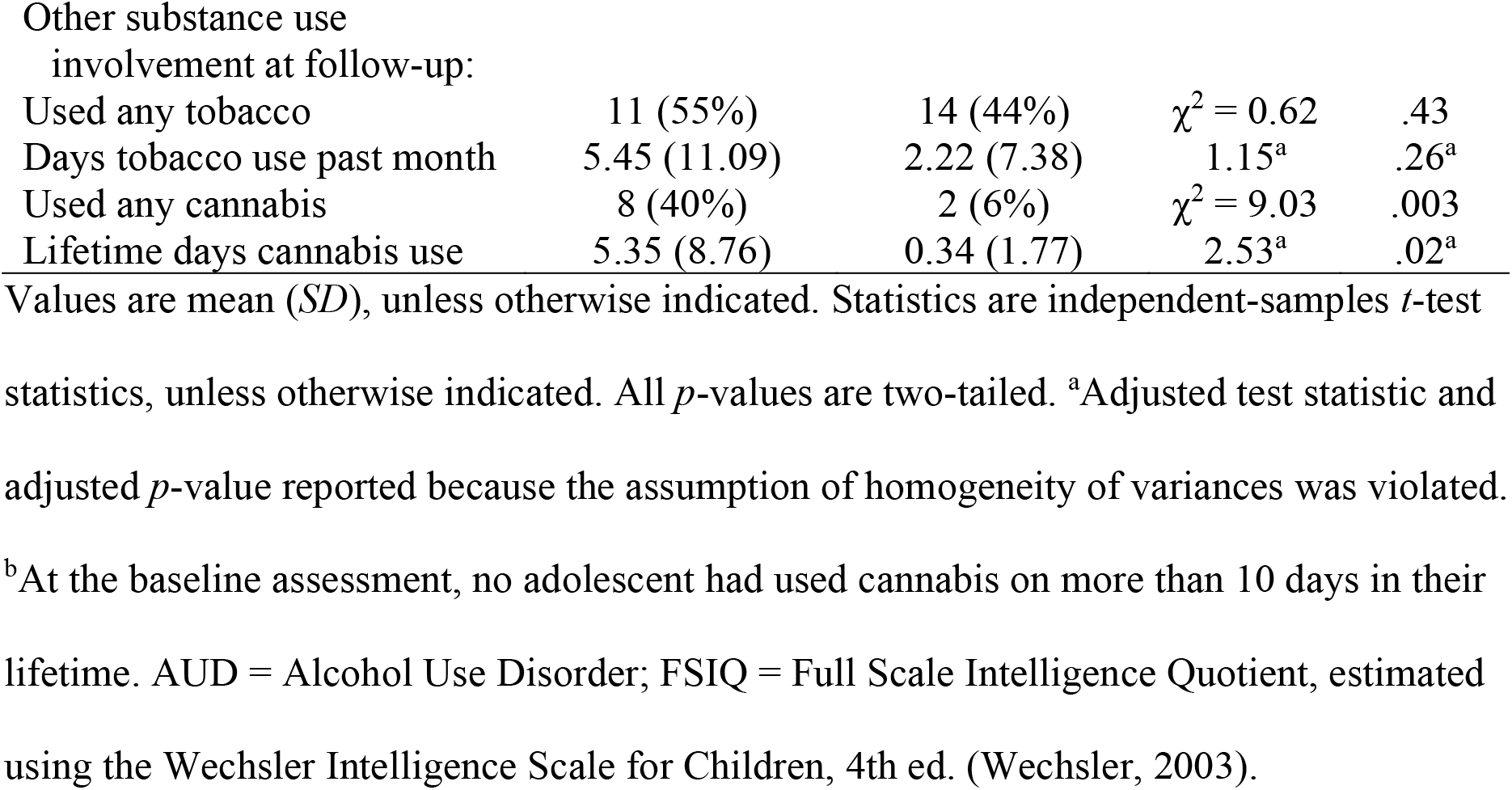
Participant Characteristics and Patterns of Alcohol and Other Drug Use.

The follow-up assessment took place approximately two years (*M* = 2.3 years ± 0.3 *SD*) after baseline when the sample had a mean age of 18.8 years (± 0.5 *SD*, age range: 17.30– 19.95 years), at which point the majority (*n* = 50, 96%) had consumed alcohol at least once in their lifetime. From among adolescents who had not consumed alcohol at least once in their lifetime when assessed at baseline (*n* = 14), one adolescent with a family history of AUD and 11 adolescents with no such family history initiated alcohol use in the follow-up period. The group of adolescents with a family history of AUD had significantly more lifetime drinking episodes at follow-up (range: 4–300 days) compared to the group of adolescents with no such family history (range: 0–100 days), and adolescents with a family history of AUD had significantly more drinking episodes in the 12 months prior to follow-up (range: 1–100 days) compared to adolescents with no such family history (range: 0–65 days). Adolescents with a family history of AUD also experienced more alcohol-related harms in the 12 months prior to follow-up (8 [40%] reported no harm; 5 [25%] reported one harm; 3 [15%] reported two harms; 2 [10%] reported three harms; 2 [10%] reported four harms) compared to adolescents with no such family history (20 [62.5%] reported no harm; 6 [19%] reported one harm; 4 [12.5%] reported two harms; 2 [6%] reported three harms), but this between-group difference was just less than conventional statistical significance (*p* = .06). Thus, our results support the notion that adolescents with a family history of AUD tend to drink more often and experience more alcohol-related harms, thereby validating our approach to determine risk status.

### Preliminary Analysis of Connectivity at Baseline

An important consideration is whether the connectomic maps of each group have comparable connection density, *κ*, calculated as the proportion of all possible connections actually present. Using the 82-node anatomical parcellation, the group average *κ* for adolescents with a family history of AUD was 74% ± 4% *SD* and for adolescents with no such family history the group average *κ* was 75% ± 4% *SD, t*(50) = 0.41, *p* = .69. With the 530-node random parcellation, the group average *κ* for adolescents with a family history of AUD was 24% ± 1% *SD* and for adolescents with no such family history the group average *κ* was 24% ± 1% *SD, t*(50) = 0.74, *p* = .46. Thus, no between-group differences were found in the overall number of inter-regional connections.

### Main Analysis of Connectivity at Baseline

We first used the NBS to identify subnetworks comprised of structural connections showing either less or greater baseline connectivity in adolescents with a family history of AUD compared to adolescents with no such family history. Using a two-tailed *t*-test statistic threshold of *p* = .05 and a component-wide FWE corrected threshold of *p* = .05, there were no significant between-group differences in baseline connectivity at either parcellation resolution using either streamline count or any of the four tensor-based measures (i.e., FA, MD, AD, and RD) as connectivity weight. Relaxing the primary threshold to *p* = .10, two-tailed, yielded the same results.

As shown in Figure 1, effect sizes (*d*) for some edges were relatively strong. However, given the number of comparisons, some edges may show large differences by chance. The lack of a statistically significant result using the NBS suggests that these large-effect edges did not aggregate into a sufficiently large subnetwork to survive correction for multiple comparisons.

**Figure 1.**
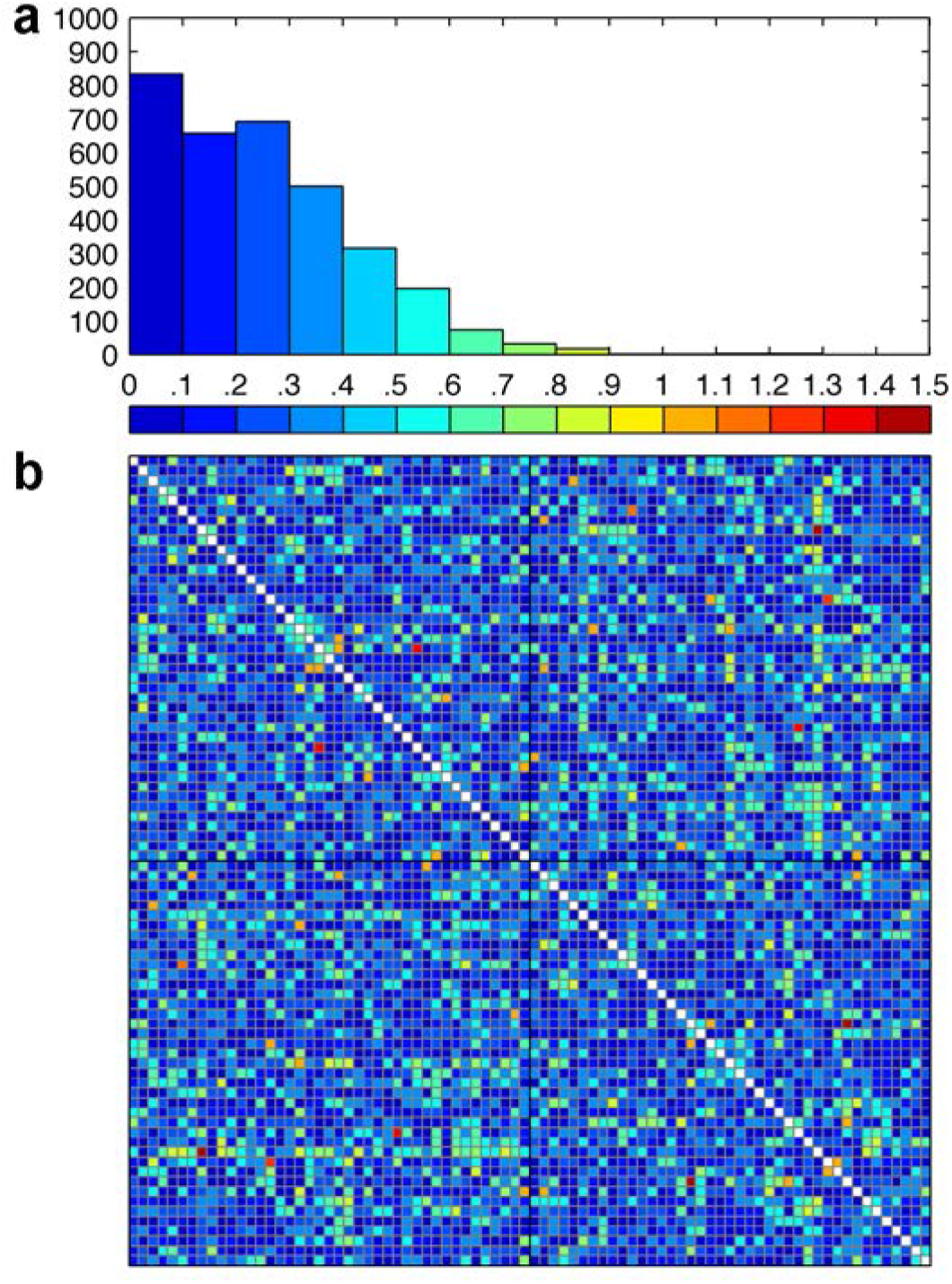
Effect sizes. ***a*** illustrates the distribution of effect sizes (*d*) for differences in streamline count-weighted structural connectivity between adolescents with and without a family history of AUD. These effect sizes populate the corresponding 82-node connectivity matrix shown in ***b***, with elements (each of which represents a unique edge, or connection) coloured according to the scheme in ***a***. Regions of interest are ordered according to hemisphere (i.e., left then right), with cortical regions first, followed by subcortical. A similar distribution of effect sizes was found for all 10 different network representations of brain connectivity.

### Correlation Analysis

The second goal of using the NBS was to examine the prospective association between baseline connectivity measures and subsequent alcohol use and its associated harms. In analyses of all 10 different network representations of brain connectivity, we found no association between baseline connectivity measures and the number of drinking episodes (days) in the 12 months prior to follow-up. There was also no association between baseline connectivity measures and the number of alcohol-related harms experienced in the 12 months prior to follow-up. These associations were also examined in adolescents with and without a family history of AUD separately and we found no evidence that baseline connectivity measures predict levels of alcohol consumption or alcohol-related harms at follow-up.

In addition to the number of drinking episodes in the 12 months prior to follow-up, alcohol use was also quantified using the difference between the number of lifetime drinking episodes at baseline and at follow-up. We found no evidence that baseline connectivity measures predict changes in lifetime drinking history between baseline and follow-up.

## Discussion

This prospective study revealed that the connectome-wide pattern of structural connectivity at an average of 16.5 years of age did not differentiate adolescents with and without a family history of AUD. Also contrary to expectations, we did not find an association between baseline structural connectivity measures and alcohol outcomes at follow-up, which took place when participants were, on average, 18.8 years of age. These findings suggest that atypical inter-regional structural connectivity may not contribute to the risk of developing alcohol use problems in this particular age group, or during this particular period of development, or alternatively that any such effects were of insufficient magnitude to be detected in connectome-wide analyses.

The previous literature includes 10 cross-sectional studies of structural and/or functional connectivity in young people with a family history of AUD (see Cservenka et al., 2015, for review). The first such study reported lower FA along several white matter tracts in 15 youths with a family history of AUD aged 11–15 years (Herting et al., 2010), suggesting that poorer white matter microstructure may be a marker of risk for alcohol use problems.

These results were further supported by a recent study in which lower FA was found in 80 youths with a family history of SUD (Acheson et al., 2014). In contrast to these findings, however, two studies examining the same sample of approximately fifty 12–14-year-olds at risk for AUD showed: (a) higher FA and AD, and lower MD and RD, in 19 different white matter tracts (Squeglia et al., 2014); and (b) lower MD and RD along white matter tracts connecting frontal and striatal regions of interest (Squeglia et al., 2015). As such, the authors suggested that the white matter of youths with a family history of AUD may be characterised by a relatively high degree of directional coherence and therefore integrity and maturity (Squeglia et al., 2014, 2015).

Herting et al. (2011) were the first to examine functional connectivity in young people with a family history of AUD, finding reduced task-based functional connectivity between bilateral anterior prefrontal cortices and contralateral cerebellar regions of interest in the same youths described in a previous study of structural connectivity (Herting et al., 2010). A further two studies examined separate samples of 12–14-year-olds at risk for AUD and both found atypical/reduced task-based functional connectivity between frontal and parietal regions of interest (Spadoni et al., 2013; Wetherill et al., 2012). These functional differences were apparent in the absence of microstructural differences in frontoparietal white matter tracts (Wetherill et al., 2012). In another study, 49 young people with a family history of AUD aged 18–22 years showed reduced task-based functional connectivity of the ventral striatum (nucleus accumbens) with precuneus and sensorimotor areas, which mediated the relationship between sensation-seeking and alcohol consumption (Weiland et al., 2013). The remaining two studies used seed-based resting-state functional connectivity methods and found group differences in the synchrony of the nucleus accumbens (Cservenka et al., 2014a) and amygdala (Cservenka et al., 2014b) with other regions of the brain between youths with and without a family history of AUD. Thus, unlike the evidence for atypical structural connectivity, which, together with our data, has been inconsistent, functional connectivity differences appear to be a more robust finding in young people with a family history of AUD.

Despite the absence of strong evidence for disturbances in structural connectivity *between* different brain regions, a large number of imaging studies have provided evidence of neural alterations *within* specific brain regions in young people with a family history of AUD (see Cservenka, 2016, for review). For example, volumetric differences between young people with and without a family history of AUD have been found in regions including orbitofrontal cortex, cingulate, hippocampus, amygdala, thalamus, and cerebellum (Benegal, Antony, Venkatasubramanian, & Jayakumar, 2007; Hanson et al., 2010; Hill et al., 2001; Hill et al., 2007b; Hill et al., 2009). In parallel studies, young people with a family history of AUD have shown atypical neural function (haemodynamic activity) when performing various cognitive tasks. Such studies have reported functional differences throughout the brain, including within frontal, parietal, temporal, and subcortical regions (Bjork, Knutson, & Hommer, 2008; Glahn, Lovallo, & Fox, 2007; Heitzeg, Nigg, Yau, Zubieta, & Zucker, 2008; Heitzeg, Nigg, Yau, Zucker, & Zubieta, 2010; Hill et al., 2007a; Rangaswamy et al., 2004; Schweinsburg et al., 2004; Silveri, Rogowska, McCaffrey, & Yurgelun-Todd, 2011).

Dysfunction within specific brain regions may alter neuronal spiking activity, potentially causing aberrant synchronisation (functional connectivity) with other areas in the absence of structural connectivity abnormalities (Fornito et al., 2015). Collectively, these findings suggest that risk for AUD in young people may be associated primarily with dysfunction of local, intra-regional circuitry, rather than macroscopic inter-regional connectivity.

Studies of adolescent substance users have found evidence of microstructural white matter abnormalities, suggesting that white matter might be impacted in the early stages of using drugs such as alcohol (Baker, Yücel, Fornito, Allen, & Lubman, 2013; Ewing et al., 2014). Further support for this notion comes from four recent longitudinal studies using diffusion-weighted MRI (Bava, Jacobus, Thayer, & Tapert, 2013; Jacobus, Squeglia, Bava, & Tapert, 2013; Jacobus, Squeglia, Infante, Bava, & Tapert, 2013; Luciana, Collins, Muetzel, & Lim, 2013). These include three studies that reported microstructural white matter alterations throughout the brain in adolescents who either initiated (Jacobus et al., 2013b) or continued (Bava et al., 2013; Jacobus et al., 2013a) heavy alcohol and cannabis use between mid and late adolescence. A fourth study followed adolescents between the ages of 14–19 (range at baseline) and 16–22 (range at follow-up) years and found evidence of microstructural alterations in those who started drinking alcohol (Luciana et al., 2013). It is important to note that the findings of these four longitudinal studies are based on only two samples, including one with a history of extensive cannabis use (Bava et al., 2013; Jacobus et al., 2013a, 2013b). Nevertheless, these findings suggest that heavy substance use in adolescence may be detrimental to white matter. Together with our evidence that long-range anatomical connections appear to be unaffected in young people with a family history of AUD, these findings suggest that the white matter abnormalities found in adolescent substance users may not reflect pre-existing differences that increase risk for alcohol and other drug problems. An important test of this hypothesis will involve longitudinal investigation of how using drugs such as alcohol and cannabis affects the connectome.

There are several important points to consider when interpreting our findings. First, our sample size was small but adequate compared to previous studies that found structural and/or functional connectivity differences (group *n*s range from 13 to 80 youths with a family history of AUD and 14 to 46 youths with no such family history). Second, the distinct characteristics of our community sample may limit the comparability of our findings to those of previous connectivity studies, most of which have examined younger adolescents who, in most cases, reported that they had never consumed alcohol or other drugs. In this regard, we also acknowledge the difficulty in dissociating risk from exposure in a sample of drinkers.

Further work in larger samples is required to determine at what stage of development any connectivity differences that may be associated with family history status are best detected and, subsequently, whether these differences confer increased risk of developing alcohol use problems. Third, connectivity measures derived from diffusion-weighted MRI can be physiologically ambiguous (Jones, Knösche, & Turner, 2013). Measures such as streamline count, FA, MD, AD, and RD are sensitive to white matter pathology, but the precise influence of specific axonal components (e.g., myelin) and architectural characteristics (e.g., geometry) on such measures is not well understood. Fourth, unlike previous connectivity studies, we tested group differences at each and every connection. We thus offer a relatively unique perspective on brain network connectivity in adolescents with a family history of AUD, but further connectome-wide studies are required to confirm our findings. Finally, although an important strength of this study is the prospective design, the follow-up period was limited to approximately two years and did not capture the entire period of highest risk for developing alcohol use problems. Thus, it is unclear whether white matter differences might predict the onset of such problems later on in emerging adulthood. Longer-term follow-up of the adolescents in this sample will provide a more complete picture of the relationship between structural connectivity and risk for alcohol use problems.

Our findings suggest a limited role for structural connectivity in contributing to the neural basis of familial/genetic risk for alcohol use problems in the examined epoch. If confirmed and extended in more powerful, prospective, longitudinal studies over longer follow-up periods, these findings will help to narrow our search for the underlying neural correlates of increased risk for the development of alcohol use problems.

## Data Availability

Data may be available on request

## Notes

This research was supported by grants from the Colonial Foundation, the Australian National Health and Medical Research Council (NHMRC; Program Grant 350241; Fellowships 1021973, 1047648, and 1007716), and the Australian Research Council (ARC; Discovery Grants DP0878136 and DP1092637; Fellowship FT130100589). Simon T. E. Baker was supported by an Australian Postgraduate Award. The authors would like to thank the adolescents and their families for participating in the Orygen Adolescent Development Study.

### Competing Interest Statement

The authors have declared no competing interest.

### Funding Statement

This research was supported by grants from the Colonial Foundation, the Australian National Health and Medical Research Council (NHMRC; Program Grant 350241; Fellowships 1021973, 1047648, and 1007716), and the Australian Research Council (ARC; Discovery Grants DP0878136 and DP1092637; Fellowship FT130100589). Simon T. E. Baker was supported by an Australian Postgraduate Award.

### Author Declarations

The University of Melbourne Human Research Ethics Committee

